# Safety and efficacy of bronchus-blocked ultrasound-guided percutaneous transthoracic needle biopsy (BUS-PTNB) for critically ill patients under invasive mechanical ventilation: a prospective, single-arm trial

**DOI:** 10.1101/2024.12.23.24319165

**Authors:** He Yu, Yun Liu, Zhen Wang, Peng Sun, Weiya Wang, Wenxi Xia, Wenli Qi, Linfei Wu, Guangdi Liu, Yaxiong Zhou, Rong Yao, Yuean Zhao, Siyu Liu, Xue Lin, Qian He, Zuoyu Liang, Wanhong Yin, Ran Zhou, Yiqun Mi, Jianjiang Luo, Hui Zhou, Huaicong Long, Zhuang Luo, Junping Fan, Charles A. Powell, Zongan Liang, Ye Wang

**Affiliations:** Department of Pulmonary and Critical Care Medicine, West China Hospital, Sichuan University, Chengdu, Sichuan, China; Department of Respiratory Care, West China Hospital, Sichuan University, Chengdu, Sichuan, China; Center of Interventional Radiology, West China Hospital, Sichuan University, Chengdu, Sichuan, China; Department of Pathology, West China Hospital, Sichuan University, Chengdu, Sichuan, China; West China Medicine Technology Transfer Center, Chengdu, Sichuan, China; Department of Emergency, West China Hospital, Sichuan University, Chengdu, Sichuan, China; Department of Anesthesiology, West China Hospital, Sichuan University, Chengdu, Sichuan, China; Department of Geriatric, Chengdu Dekang Hospital, Chengdu, Sichuan, China; Department of Critical Care Medicine, West China Hospital, Sichuan University, Chengdu, Sichuan, China; Hangzhou Matridx Biotechnology Co., Ltd, Hangzhou, Zhejiang, China; Department of Respiratory Medicine, Traditional Chinese Medicine Hospital, Xinjiang Medical University, Urumqi, Xinjiang, China; Department of Respiratory and Critical Care Medicine, Affiliated Hospital & Clinical College of Chengdu University, Chengdu, Sichuan, China; Department of Geriatric Intensive Care Unit, Sichuan Provincial People’s Hospital, University of Electronic Science and Technology of China, Chengdu, Sichuan, China; Department of Respiratory and Critical Care Medicine, First Affiliated Hospital of Kunming Medical University, Kunming, Yunnan, China; Department of Pulmonary and Critical Care Medicine, Peking Union Medical College Hospital, Chinese Academy of Medical Sciences & Peking Union Medical College, Beijing, China; Division of Pulmonary, Critical Care and Sleep Medicine, Icahn School of Medicine at Mount Sinai, New York, NY, USA; Precision Medicine Key Laboratory of Sichuan Province, West China Hospital, Sichuan university, Chengdu, Sichuan, China

**Keywords:** Biopsy, Needle, Critical Care, Respiratory Infection, Assisted Ventilation

## Abstract

**Objectives:** Conventional lung biopsy for critically ill patients who under invasive mechanical ventilation (IMV) is limited due to high risks of procedure-related complications. We developed a novel technique named bronchus-blocked ultrasound-guided percutaneous transthoracic needle biopsy (BUS-PTNB) to mitigate the risks of biopsy under IMV, but its safety and efficacy have not been prospectively evaluated.

**Methods:** In this prospective, single-arm trial (Chictr.org, ChiCTR2100054047), invasively ventilated patients with undiagnosed lung opacities were screened and underwent BUS-PTNB after enrollment. The peri-operative conditions, severity of complications, pathological findings, and tissue-based metagenomic next-generation sequencing (mNGS) results were systemically evaluated.

**Results:** A total of 22 critically ill patients (18 men, mean age, 64.2 years [SD 11.7], APACHE II score, 27.0 [SD 5.6], PaO_2_/FiO_2_ ratio, 120.5 [IQR 91.9–169.1]) under IMV were enrolled in the study. Throughout the procedure, there were no significant changes in respiratory rate or PaO_2_. Biopsy-related complications occurred in 3 patients (13.6%), including pneumothorax (n=1, 4.5%), intrabronchial hemorrhage (n=2, 9.1%), and hemothorax (n=3, 13.6%). Notably, one patient (P10) required a blood transfusion due to hemothorax, which was classified as a severe complication (1/22, 4.5%). Satisfactory biopsy samples were obtained from 21 patients (95.5%) for pathological study and from all 22 patients (100%) for mNGS.

**Conclusions:** **The novel** BUS-PTNB is a promising bedside biopsy technique for ICU patients under IMV with acceptable complication risk. This technique may prove instrumental in advancing pathological studies of severe lung diseases.

**Key points and Clinical Relevance Statement:** *Question:* We developed bronchus-blocked ultrasound-guided percutaneous transthoracic needle biopsy (BUS-PTNB) for patients under invasive mechanical ventilation, but its safety and efficacy have not been prospectively evaluated.

*Findings:* Patients’ vital signs showed no significant fluctuations. Biopsy-related complications occurred in 3 patients out of 22. Satisfactory samples were obtained for pathological study and mNGS.

*Clinical Relevance:* BUS-PTNB is a promising bedside biopsy technique for ICU patients under IMV with acceptable complication risk. This technique may prove instrumental in advancing pathological studies of severe lung diseases.

## Introduction

Lung opacities with respiratory failure of various etiologies can progress rapidly[1]. 68.8% ICU patients require mechanical ventilation because of acute respiratory failure[2]. Timely diagnosis of the underlying etiologies and identification of the pathogens are crucial for personalizing treatments and improving outcomes[3, 4]. Although significant progress has been achieved in novel bronchoalveolar lavage fluid (BALF) assays, blood testing, and imaging examinations[5–8], pathological evaluation remains the diagnostic gold standard for malignancies, which is also valuable for informing etiological diagnosis and pathogen identification in inflammatory and interstitial lung diseases[9–12].

Obtaining lung specimens in ICU is challenging, especially in patients under invasive mechanical ventilation (IMV). The high risks of bleeding and pneumothorax of biopsy associated with positive pressure ventilation must be balanced against their potential benefits[13–17]. Conventional open lung biopsy or video-assisted thoracic surgery (VATS) are often used in the past, but only for highly selective patients[18–21]. Other minimally invasive techniques, such as transbronchial lung biopsy (TBLB), percutaneous transthoracic needle biopsy (PTNB), and transbronchial cryoprobe lung biopsy (TCLB), may obtain sufficient specimens for pathological studies with acceptable safety[22–26]. However, these approaches would be less effective for accessing peripheral lung lesions and the acquired sample may suffer a risk of contamination by airway-colonizing organisms[13, 27]. This gap in availability of a widely acceptable biopsy technique for critically ill patients under IMV impacts pathological diagnostics and translational research into severe lung diseases.

To address this gap, we developed a novel biopsy technique named bronchus-blocked ultrasound-guided percutaneous transthoracic needle biopsy (BUS-PTNB)[28]. Despite acceptable complications reported in retrospective literature, the reality is that the clinicians tend to avoid using these biopsy techniques because of their uncertainty of potentially mortal complications. Hence, we experimented with a physical safeguard in the BUS-PTNB procedure, an inflatable balloon for blocking the bronchus during specimen collection. According to our earlier feasibility trial, the blocker could effectively prevent blood from flowing into the proximal airway and air leakage into the thoracic cavity. Meanwhile, sufficient tissue specimens could be attained for subsequent pathological diagnostics. We described the feasibility and workflow of BUS-PTNB in a previous report on four pilot cases of severe lung diseases under IMV[28]. However, the overall safety and efficacy profile of the technique remained uninvestigated. Therefore, we conducted this prospective, single-arm clinical trial in a larger cohort to validate the safety and efficacy of BUS-PTNB.

## Materials and Methods

### Ethical Statement

All procedures were in line with the ethical standards of the IRB and the Helsinki Declaration. Written informed consents were obtained from every participant in accordance with standard procedures. Ethical approval was obtained from the Institutional Review Board of West China Hospital of Sichuan University (approval number: 2021-1250). The study has been declared on Chinese Clinical Trial Registry (ChiCTR2100054047).

### Study Design and Participants

This was an investigator-initiated, prospective, single-arm clinical trial conducted at a medical intensive care unit (MICU) in West China Hospital, Sichuan University between December 2021 and July 2022. We screened candidate patients who were admitted to the emergency intensive care unit (EICU) or the MICU and invasive-mechanically ventilated for respiratory failure with lung opacities of unknown cause. Patients were included if aged 18-80 years; having been placed on IMV; chest computed tomography (CT) indicated peripheral/sub-pleural opacities (including but not limited to consolidation, plaques, nodules, and masses) or diffuse interstitial changes of unknown cause; or existing clinical data failed to explain the pulmonary disease progression. The exclusion criteria included arterial oxygen partial pressure (PaO_2_) <60 mmHg under optimal mechanical ventilation, platelet count <50 ×10^9^/L, an international normalized ratio >1.5, or a fibrinogen level <1 g/L; pregnancy; informed consent not obtained from patient or family; notable disease alleviation; or any other paroxysmal comorbid condition which could cause sudden deterioration or compromise the procedure, such as a recent episode of malignant arrhythmia or epilepsy.

### BUS-PTNB Procedure

The BUS-PTNB procedures were performed at bedside in the MICU by a 5-member operation team, including a critical care pulmonologist, an interventional pulmonologist, two respiratory therapists, and a nurse. In our previous article we described the 7-step EPUBNOW workflow, *i.e.* evaluation, preparation, ultrasound location, bronchus blocking, needle biopsy, observation, and withdrawal of blocker[28]. In this trial, we added needle tract plugging after sampling to the needle biopsy step for improving prevention of hemothorax (Supplementary Figure 2).

The workflow and role-based task assignments was as follows (Supplementary Figure 1). The critical care pulmonologist evaluated the patient and assessed the necessity of biopsy. The interventional pulmonologist evaluated the feasibility of procedure, intended lobe, and potential risks. The respiratory therapist evaluated the ventilation, oxygenation, and airway conditions. The intended lobe and insertion site were evaluated with latest CT and real-time sonography. During preparation, the patient’s Richmond Agitation and Sedation Scale (RASS) was kept at -4. Then, an endobronchial blocker was inserted by bronchoscopy and placed in the intended lobar bronchus. If the SpO_2_ was maintained >90% for 5 min after bronchus blocking, needle biopsy was initiated under the supervision of intrabronchial view with the bronchoscope. In case intrabronchial hemorrhage was detected, the location and inflation of the blocker were adjusted to prevent blood flow into the airway. After sampling, 2-3 ml saline mixed with gelatin sponge and thrombin was injected into the coaxial needle to plug the needle tract when needed. After biopsy, the blocker was fixed, and the patients’ condition were observed for at least 2 hours. The blockers were withdrawn under bronchoscopy supervision, and the blood clots if any in the distal airway were aspirated.

### Outcomes

The primary endpoint was the incidence and severity of procedure-associated complications. A procedure-associated complication was defined as an adverse condition resulted from the BUS-PTNB procedure within 48 hours post procedure, including pneumothorax, intrabronchial hemorrhage, and hemothorax. Sonography was routinely performed 2, 24, and 48 hours after procedure to detect hemothorax and pneumothorax. Chest X-ray was performed around 48 hours after procedure and then repeated *ad hoc*. Intrabronchial hemorrhage was evaluated by bronchoscopy during and after biopsy. Mild (Grade 1) and moderate (Grade 2) complications had no or little influence on a patient’s overall condition and outcome, where a Grade 1 complication required no inventions, and a Grade 2 complication only required simple management. A severe (Grade 3) complication caused adverse impact on patient condition that required surgical intervention(s) and/or blood transfusion. A Grade 3 complication might affect patient outcome and prolong hospital stay. A life-threatening (Grade 4) complication caused persistent, severe changes in a patient’s vital signs and required emergent management, which could result in organ dysfunction and a poor outcome. In this trial, the procedure was considered safe if the incidence of Grade 3 complication was <15% and Grade 4 complication < 5%.

The secondary endpoint was efficacy of procedure (success rate of tissue collection sufficient for pathological studies). The fresh lung tissues underwent mNGS to detect pathogens, if necessary.

### Statistical Analysis

The sample size was estimated using the single-arm objective performance criteria based on the acceptable rate of severe complication.

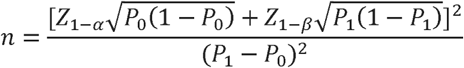

According to previous reports, 73.0-76.9% pathological results had strong clinical significance and 84.0-95.3% provided important information for etiological diagnoses[29–32]. After balancing the above benefit rates, we estimated our sample size using an assumed 15% acceptable incidence of severe complication. As calculated, 22 patients were needed for 80% power, at an alpha level of 2.5%.

The trial would be discontinued in event of > 3 cases of Grade 3 complication, > 1 case of Grade 4 complication, or 1 instant procedure-related death.

Continuous variables were presented as means with standard deviations (SD, normal distribution) or medians with interquartile ranges (IQR, skewed distribution). *t*-test and Wilcoxon test were calculated for data comparison. *p* <0.05 was statistically significant.

## Results

### Patient Characteristics

105 consecutive patients with undiagnosed lung opacities were screened between December 2021 and July 2022. Finally, 22 patients were enrolled and underwent BUS-PTNB (Figure 1). Table 1 details their baseline conditions. 9 (40.9%) patients had acute respiratory distress syndrome (ARDS) and 11 (50.0%) were comorbid with septic shock. Notably, 3 (13.6%) patients had anatomically altered airways because they had undergone lobectomy for early-stage lung cancers. The mean APACHE II score was 27.0 (SD 5.6). The median PaO_2_/FiO_2_ ratio was 120.5 (IQR 91.5-169.3). For mechanical ventilation, tidal volume was set at a median of 410 ml (IQR 400-445) and median positive end expiratory pressure (PEEP) at 6.0 cmH_2_O (IQR 5.0-9.8). 16 (72.7%) patients had bilateral/diffuse lung opacities and 6 (27.3%) had regional lung opacities, including consolidations (10, 45.5%), interstitial opacities (7, 31.8%), ground glass opacities (3, 13.6%), nodules (1, 4.5%), and mass (1, 4.5%) (Supplementary Figure 3). The median duration of invasive ventilation was 64.0 hours before BUS-PTNB.

**Figure 1.**
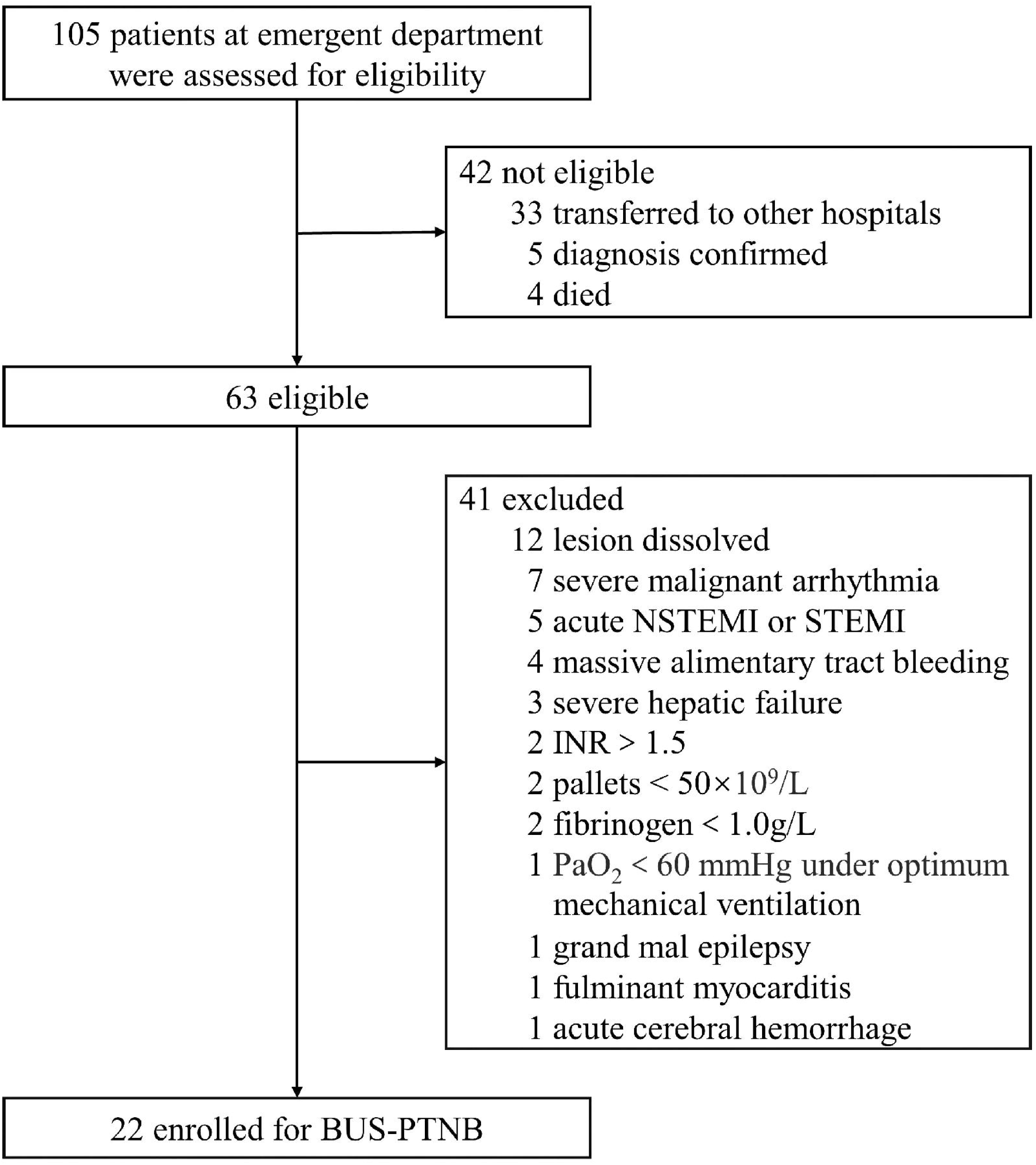
Flowchart of the study. NSTEMI = non-ST-segment elevation myocardial infarction, STEMI = ST-segment elevation myocardial infarction, INR = international normalized ratio, PaO_2_ = arterial oxygen pressure, BUS-PTNB = bronchus-blocked ultrasound-guided percutaneous transthoracic needle biopsy

**Table 1.** Baseline Characteristics.

| Variables | Value (N=22) |  |
| --- | --- | --- |
| Demographics |  |  |
| Age, yr, mean (SD) | 64.2 | (11.7) |
| Male, *n* (%) | 18 | (81.8) |
| Female, *n* (%) | 4 | (18.2) |
| Baseline conditions, *n* (%) |  |  |
| Septic shock | 11 | (50.0) |
| ARDS | 9 | (40.9) |
| Pneumonia | 6 | (27.3) |
| Interstitial lung disease | 5 | (22.7) |
| Post-lobectomy | 3 | (13.6) |
| Assessment and measurement |  |  |
| APACHE II score, mean (SD) | 27 | (5.6) |
| Total SOFA score, mean (SD) | 9.7 | (3.4) |
| PaO<sub>2</sub>/FiO<sub>2</sub>, mmHg, median (IQR) | 120.5 | (91.5 – 169.3) |
| PEEP, cmH<sub>2</sub>O, median (IQR) | 6.0 | (5.0–9.8) |
| RR, breath per minute, mean (SD) | 24 | (6) |
| Tidal volume, ml, median (IQR) | 410.0 | (400.0 – 445.0) |
| CT findings, *n* (%) |  |  |
| Diffused/Bilateral | 16 | (72.7) |
| Regional | 6 | (27.3) |
| Consolidation | 10 | (45.5) |
| Interstitial change | 7 | (31.8) |
| Patchy shadow | 3 | (13.6) |
| Nodule | 1 | (4.5) |
| Mass | 1 | (4.5) |
| Comorbid conditions, *n* (%) |  |  |
| Hepatic disease | 11 | (50.0) |
| Hypertension | 9 | (40.9) |
| Renal failure | 8 | (36.4) |
| DVT | 7 | (31.8) |
| Malignant tumor | 7 | (31.8) |
| Immunocompromise | 6 | (27.3) |
| Diabetes mellitus | 5 | (22.7) |
| CHD | 4 | (18.2) |
| COPD | 2 | (9.1) |
| Duration of invasive mechanical ventilation, hours, median (IQR) | 64 | (41.3 – 183.5) |

ARDS = acute respiratory distress syndrome, APACHE II = Acute Physiology and Chronic Health Evaluation II, Total SOFA = Total Sequential Organ Failure Assessment, PaO<sub>2</sub>/FiO<sub>2</sub> = arterial pressure of oxygen/inspiratory fraction of oxygen, PaO<sub>2</sub> = arterial pressure of oxygen, FiO<sub>2</sub> = inspiratory fraction of oxygen, PEEP = positive end expiratory pressure, RR = respiratory rate, DVT = deep venous thrombosis, CHD = coronary heart disease, COPD = chronic obstructive pulmonary disease.

### BUS-PTNB Procedure

Table 2 details the BUS-PTNB procedure. The S9 segment in the lower lobes (RL/S9, LL/S9) was the biopsy site of choice in 12 (54.6%) patients and the upper lobes (RU/S2, LU/S3) in 2 (9.1%) patients. Accordingly, we blocked the right lower lobar bronchi in 12 (54.5%) patients, the left lower lobar bronchi in 7 (31.8%) patients, and the right intermediate bronchi in 4 (18.2%) patients. Blocker placement took 12.7 min (SD 6.1), needle biopsy 14.0 min (IQR 12.0-16.8), and bronchus blockage lasted for 222.0 min (IQR 148.8-258.5), respectively. Needle tract plugging with a mixture of gelatin sponge and thrombin was applied in the 12 (54.5%) procedures.

**Table 2.**
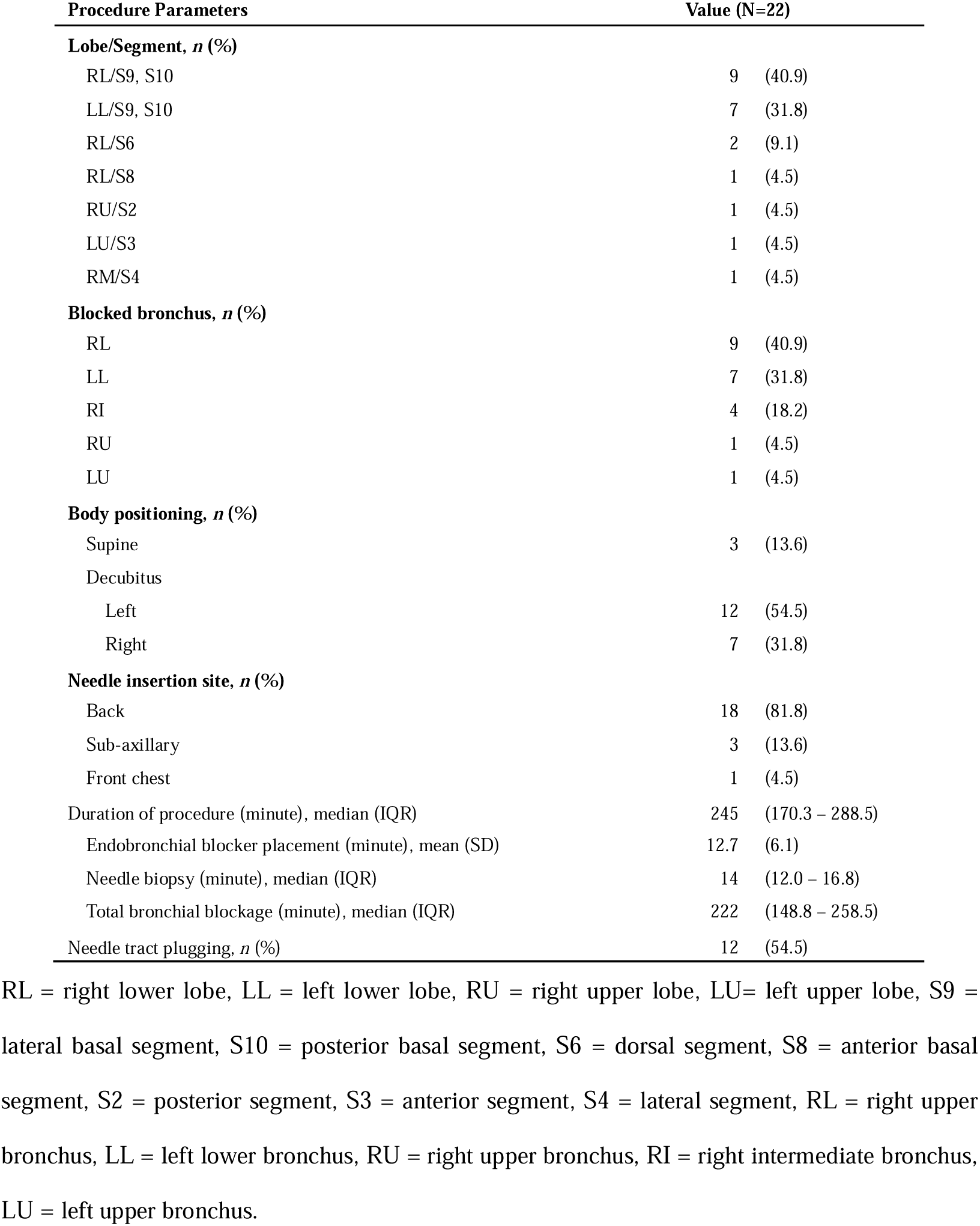
Procedure Information.

### Peri-procedural Conditions

Figure 2 illustrates the patients’ peri-operative conditions and ventilator monitoring values. Stimulated by the endobronchial blocker, patient might cough during blocker placement, for which small amount of *i.v.* sedatives and analgesics was given. Lowering of blood pressure (BP) was noted when repositioning or on use of sedatives during procedure. Norepinephrine was administered to maintain BP when appropriate. The peak and mean airway pressures increased during bronchoscopy after blocker placement, which returned to the baseline level after the bronchoscope was withdrawn during post-procedure observation while the blocker still in place. This suggested that the increased airway pressure might be resulted from the use of the bronchoscope instead of the presence of the endobronchial blocker.

**Figure 2.**
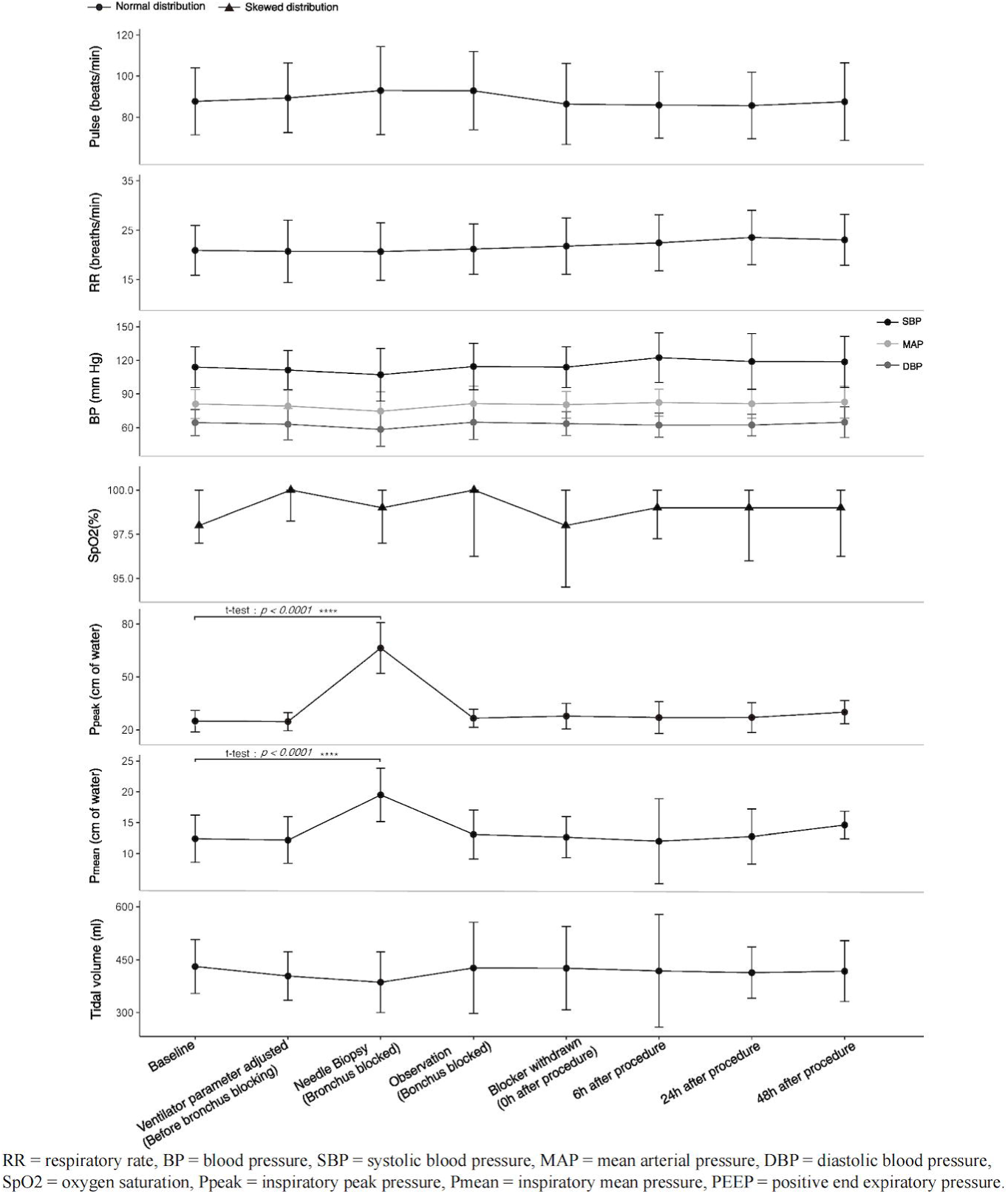
Peri-operative conditions. The peak and mean pressures were significantly higher when inserting bronchoscope into the airway. The other vital signs and ventilation status did not change significantly throughout the procedures. Error bars represent SD or IQR. **** *p* < 0.0001.

### Complications

Procedure-associated complications occurred in 3 (13.6%) patients (Table 3), including 2 (9.1%) patients with Grade 2 complications and 1 (4.5%) patient with Grade 3 complications. There was no Grade 4 complication.

**Table 3.** Complications.

| Patient | Complication | Severity Grade | Possible Cause | Management |
| --- | --- | --- | --- | --- |
| P8 | PNX | 2 | Incomplete blocking | Adjust blocker's position, re-inflate balloon |
|  | HT | 1 | Needle tract bleeding | Closed thoracic drainage |
|  | IH | 1 | Incomplete blocking | Adjust blocker's position, re-inflate balloon |
| P9 | HT | 2 | Needle tract bleeding | Closed thoracic drainage |
| P10 | IH | 2 | Incomplete blocking | Adjust blocker's position, re-inflate balloon |
|  | HT | 3 | Needle tract bleeding | Closed thoracic drainage, blood transfusion |

PNX = pneumothorax, HT = hemothorax, IH = intrabronchial hemorrhage.

P8 had a relatively high opening of dorsal segmental bronchus, so the middle lobe was partially uncovered. Pneumothorax (Grade 2 complication) and intrabronchial hemorrhage (Grade 1) occurred during sampling, which was immediately managed by adjusting the endobronchial blocker to the intermediate bronchus. A thoracic tube was placed for drainage, in which blood was seen, indicating possible hemothorax (Grade 1).

Sonography after procedure detected increased fluid in the thoracic cavity of P9. Possible intrathoracic bleeding (Grade 2) was considered and a chest tube was placed.

P10 also had a high opening of the dorsal segmental bronchus, making it hard to only block the lower lobe bronchus. Intrabronchial hemorrhage (Grade 2) from the dorsal segment was detected and managed by adjusting the blocker to the intermediate bronchus during sampling. Post-procedure intrathoracic bleeding (Grade 3) was detected, for which blood transfusion was given.

Needle tract plugging, achieved by injecting a mixture of gelatin sponge and thrombin in normal saline into the needle tract via a coaxial needle, was employed in 12 patients, and no instances of hemothorax were observed.

### Efficacy and Clinical Outcomes

Tissue specimens obtained were satisfactory for pathological studies from 21 (95.5%) patients and mNGS testing from 22 (100%) patients (Table 4). The main pathological diagnoses included organizing pneumonia (9, 40.9%), diffuse alveolar damage (DAD) (3, 13.6%), malignancies (3, 13.6%), neutrophil infiltration (3, 13.6%), acute fibrotic and organizing pneumonia (AFOP) (1, 4.5%), invasive aspergillosis (1, 4.5%), and non-specific inflammation (1, 4.5%) (Supplementary Figure 3).

**Table 4.** Biopsy Results and Clinical Outcomes.

| **Results and Outcomes** | **N=22** |
| --- | --- |
| **Pathological findings, *n* (%)** |  |
| Organizing pneumonia | 9 (40.9) |
| DAD | 3 (13.6) |
| Neutrophil infiltration | 3 (13.6) |
| Malignancy | 3 (13.6) |
| Invasive aspergillosis | 1 (4.5) |
| AFOP | 1 (4.5) |
| Non-specific chronic inflammation | 1 (4.5) |
| No intended lesion | 1 (4.5) |
| **Tissue mNGS, *n* (%)** |  |
| Positive results | 13 (59.1) |
| *Klebsiella pneumoniae* | 3 (13.6) |
| *Acinetobacter baumannii* | 2 (9.1) |
| *Epstein-Barr virus* | 2 (9.1) |
| *Staphylococcus aureus* & *Klebsiella pneumoniae* | 1 (4.5) |
| *Escherichia coli* & *Klebsiella pneumoniae* | 1 (4.5) |
| *Pneumocystis jirovecii* | 1 (4.5) |
| *Aspergillus flavus* | 1 (4.5) |
| *Influenza B virus* | 1 (4.5) |
| *Legionella* | 1 (4.5) |
| Negative results | 9 (40.9) |
| **Therapeutic changes, *n* (%)** |  |
| Adding corticosteroids | 8 (36.4) |
| Adding antibiotics | 6 (27.3) |
| Discontinuing or de-escalating antibiotics | 6 (27.3) |
| Non-specific findings without therapeutic changes | 2 (9.1) |
| **Outcomes, *n* (%)** |  |
| Improved and discharged | 11 (50.0) |
| Died or discharged (abandoned treatment) | 11 (50.0) |

DAD = diffuse alveolar damage, AFOP = acute fibrinous and organizing pneumonia, mNGS = metagenomic next-generation sequencing

Tissue mNGS detected definitive pathogens in 13 (59.1%) patients (including P2). Except 2 results reporting *Epstein-Barr virus*, 11 (50.0%) results were important for guiding personalized anti-infectious treatments. Antibiotics were discontinued or de-escalated in 6 (27.3%) patients because of bacteria-negative reports.

The pathological studies provided important information for the subsequent therapeutic decisions in 20 (90.9%) patients, including 4 (18.2%) diagnoses (3 malignancies, 1 AFOP) that relied on the pathological findings exclusively. 11 (50.0%) patients were discharged after improvement and 11 (50%) patients were discharged automatically or deceased.

## Discussion

We investigated the safety and efficacy of BUS-PTNB, a novel biopsy technique, in critically ill patients under IMV. The overall incidence of complication was 13.6% (3/22), with only one case of severe complication (hemothorax). 95.5% (21/22) patients’ biopsies yielded satisfactory specimens for both pathological studies and mNGS testing. This demonstrates BUS-PTNB as a promising lung biopsy technique applicable for invasively ventilated patients with severe lung diseases.

The key innovation of BUS-PTNB is the introduction of an endobronchial blocker in the intended lobar bronchus for preventing possible pneumothorax and intrabronchial hemorrhage with conventional cutting needle biopsy. As expected, the incidence of pneumothorax was as low as 4.5% (1/22) using BUS-PTNB, compared with 5.5-26.6% using conventional techniques. With the bronchus blockage, intrabronchial bleeding during or after lung biopsy occurred only in 2 (9.1%) of our patients, considerably lower than other reports (4-27%)[32, 33].

According to our analyses of the two intrabronchial hemorrhage and pneumothorax cases, the complications were resulted from incomplete blockage of the right lower lobar bronchi. Because of the patients’ congenital variations and lobectomy, their bronchus openings of the right dorsal segments were high, at the same level as the middle lobe bronchi. This made it infeasible to completely block the lower lobes. We attempted to avoid covering the middle lobar bronchi by not fully inflating the blocker. This led to subsequent pneumothorax and intrabronchial bleeding. The bleeding was successfully managed by immediately pulling the blocker backwards to the intermediate bronchus. For the patients after, their anatomic structures of the right bronchi were carefully evaluated before procedure, which should be made part of the evaluation step in the standard BUS-PTNB workflow in the future. If a patient had very high opening of the right dorsal segmental bronchus, the intermediate bronchus would be preferable for blocking.

The patients in our trial had a higher mean SOFA score (9.7) compared with those in previous studies (6.5-7.0), indicating that our patients were in worse conditions at the time of biopsy[34, 35].A concern with bronchus blockage was possible influences on oxygenation status and vital signs. In our trial, the SpO_2_ levels did not decrease after bronchus blockage in most patients. A possible explanation is that FiO_2_ was adjusted to 100% and PEEP was maintained at the baseline level during procedure. The bronchus blockage allowed higher intro-procedure PEEP without increasing the risk of pneumothorax. Body position change and use of sedatives during procedure seemed to affect the patients’ BP levels. Vasopressors were administered when the mean arterial blood pressure (mABP) was <65 mmHg. The other vital signs did not fluctuate significantly.

The use of an endobronchial blocker significantly reduced the incidences of pneumothorax and intrabronchial bleeding. However, intrathoracic bleeding was observed in 3 of the first 10 patients (P8, P9, P10), including one case classified as a Grade 3 (severe) complication. Notably, the application of needle tract plugging effectively prevented hemothorax. Furthermore, during the withdrawal of the blocker, we noted reduced clot formation in the blocked bronchus. Our findings suggest that the BUS-PTNB technique substantially enhances safety as a novel biopsy procedure for invasively ventilated patients in ICU settings, potentially fostering clinician confidence and promoting increased utilization of biopsy for pathological and other diagnostic purposes in critically ill patients under IMV.

The success rate of BUS-PTNB for tissue specimen collection is excellent at 95.5% (21/22). According to our retrospective analysis of the only failed case (P2), the main cause of failure in sampling is inappropriate body positioning and overreliance on ultrasound guidance. It was difficult to collect specimens in RL/S9 in the supine position. As a result, the biopsy operator inserted the needle from an incorrect angle and pushed it to a depth very close to the diaphragm.

This resulted in failure to collect the intended tissues. In resolving it, we placed all patients after P2 in the decubitus position for sampling in S9/S10. Depths and directions of insertion were checked by pre-procedural CT and monitored using real-time ultrasound imaging.

Subsequent pathological studies provided therapeutic-instructive information in 20 (90.9%) patients, higher than reported in previous retrospective studies, suggesting that BUS-PTNB can be instrumental for supporting steroid treatment, infection detection, and tumor diagnosis[29–32]. Additionally, tissue-based mNGS testing identified single or multiple pathogens in 11 (50%) cases, which were consistent with the conventional culture findings and clinical manifestations and may be a useful reference for selecting anti-infection regimens. Notably, negative results of tissue-based mNGS might indicate absence of ongoing bacterial infections. Some microbes, such as *Candida*, *Acinetobacter Baumannii*, and *Mycobacteria Abscess* were detected by airway aspirate culture or BALF mNGS but not in the biopsy tissues, suggesting possible colonization in the airway instead of infection. Eventually, half of the patients were discharged after improvement and the other half experienced poor prognosis. In summary, the pathological studies and mNGS testing using the lung tissues collected by BUS-PTNB are informative for more precise diagnostic and therapeutic decisions and bridging the gap in knowledge about the physiopathology of severe lung diseases.

This study had severe limitations. First, this was a single-arm trial. Needle biopsy without bronchial blockage might have extraordinarily high risk in critically ill patients. We didn’t set the control group for safety reasons and ethical concerns. Second, it has a relatively small sample size. We thought this novel procedure might have some potential defects and needed to be refined during the trial, so we proposed a minimum sample size based on the primary goal of observing the procedure safety. We planned a larger, prospective, multi-center study to compare severe lung biopsy techniques for critically ill patients in near future. Third, we excluded the patients with severe cardiac or cerebral diseases. We thought some paroxysmal comorbid condition could cause sudden deterioration during the procedures and would be difficult to differentiate them from procedure-related complications.

BUS-PTNB is a promising bedside biopsy technique for ICU patients under IMV with acceptable complication risk, providing sufficient specimen for pathological studies and mNGS testing. It may address the gap in biopsy methodology in the intensive care settings and yield satisfactory specimens to support diagnostics and therapeutic decision-making.

## Data Availability

All data produced in the present study are available upon reasonable request to the authors

## List of abbreviations

BUS-PTNB: Bronchus-blocked ultrasound-guided percutaneous transthoracic needle biopsy
IMV: Invasive mechanical ventilation
mNGS: Metagenomic next-generation sequencing
BALF: Bronchoalveolar lavage fluid
VATS: Video-assisted thoracic surgery
TBLB: Transbronchial lung biopsy
TCLB: Transbronchial cryoprobe lung biopsy
MICU: Medical intensive care unit
EICU: Emergency intensive care unit
CT: Computed tomography
PaO2: Arterial oxygen partial pressure
RASS: Richmond Agitation and Sedation Scale
SD: Standard deviations
IQR: Interquartile ranges
ARDS: Acute respiratory distress syndrome
PEEP: Positive end expiratory pressure
BP: Blood pressure
DAD: Diffuse alveolar damage
AFOP: Acute fibrotic and organizing pneumonia
mABP: Mean arterial blood pressure

## Notes

### Competing Interest Statement

The authors have declared no competing interest.

### Clinical Trial

ChiCTR2100054047

### Funding Statement

The study was funded by 135 Project for Disciplines of Excellence, Clinical Research Incubation; 135 Project for Disciplines of Excellence, West China Hospital, Sichuan University (grant No. 2022HXFH005, ZYJC21032).

### Author Declarations

IRB of West China Hospital gave ethical approval for this work. The trial protocol was approved by the West China Hospital Ethics Board (approval # 2021-1250) on 2021.11.19 and registered with the Chinese Clinical Trial Registry (registration # ChiCTR2100054047) on 2021.12.07.

## References

1. Bellani G, Laffey JG, Pham T et al (2016) Epidemiology, Patterns of Care, and Mortality for Patients With Acute Respiratory Distress Syndrome in Intensive Care Units in 50 Countries. JAMA 315(8):788-800

2. Esteban A, Anzueto A, Frutos F et al (2002) Characteristics and outcomes in adult patients receiving mechanical ventilation: a 28-day international study. JAMA 287(3):345–355

3. Beitler JR, Thompson BT, Baron RM et al (2022) Advancing precision medicine for acute respiratory distress syndrome. The Lancet Respiratory Medicine 10(1):107–120

4. Matthay MA, Arabi YM, Siegel ER et al (2020) Phenotypes and personalized medicine in the acute respiratory distress syndrome. Intensive Care Med 46(12):2136–2152

5. Papazian L, Calfee CS, Chiumello D et al (2016) Diagnostic workup for ARDS patients. Intensive Care Med 42(5):674–685

6. Kyo M, Hosokawa K, Ohshimo S, Kida Y, Tanabe Y Shime N (2020) Prognosis of pathogen-proven acute respiratory distress syndrome diagnosed from a protocol that includes bronchoalveolar lavage: a retrospective observational study. J Intensive Care 8:54

7. Kao KC, Chiu LC, Hung CY et al (2017) Coinfection and Mortality in Pneumonia-Related Acute Respiratory Distress Syndrome Patients with Bronchoalveolar Lavage: A Prospective Observational Study. Shock 47(5):615–620

8. Gu W, Deng X, Lee M et al (2021) Rapid pathogen detection by metagenomic next-generation sequencing of infected body fluids. Nat Med 27(1):115–124

9. Trahan S, Hanak V, Ryu JH Myers JL (2008) Role of surgical lung biopsy in separating chronic hypersensitivity pneumonia from usual interstitial pneumonia/idiopathic pulmonary fibrosis: analysis of 31 biopsies from 15 patients. Chest 134(1):126–132

10. Li H, Gao H, Meng H et al (2018) Detection of Pulmonary Infectious Pathogens From Lung Biopsy Tissues by Metagenomic Next-Generation Sequencing. Front Cell Infect Microbiol 8:205

11. Guo Y, Li H, Chen H et al (2021) Metagenomic next-generation sequencing to identify pathogens and cancer in lung biopsy tissue. EBioMedicine 73:103639

12. Aublanc M, Perinel S Guerin C (2017) Acute respiratory distress syndrome mimics: the role of lung biopsy. Curr Opin Crit Care 23(1):24–29

13. Malhotra A Patel S (2006) Lung biopsy in ARDS: is it worth the risk? Crit Care 10(4):160

14. Pincus PS, Kallenbach JM, Hurwitz MD et al (1987) Transbronchial biopsy during mechanical ventilation. Crit Care Med 15(12):1136–1139

15. Bulpa PA, Dive AM, Mertens L et al (2003) Combined bronchoalveolar lavage and transbronchial lung biopsy: safety and yield in ventilated patients. Eur Respir J 21(3):489–494

16. Ghiani A Neurohr C (2021) Diagnostic yield, safety, and impact of transbronchial lung biopsy in mechanically ventilated, critically ill patients: a retrospective study. BMC Pulm Med 21(1):15

17. Palakshappa JA Meyer NJ (2015) Which patients with ARDS benefit from lung biopsy? Chest 148(4):1073-1082

18. Papazian L, Thomas P, Bregeon F et al (1998) Open-lung biopsy in patients with acute respiratory distress syndrome. Anesthesiology 88(4):935–944

19. Bensard DD, McIntyre RC, Jr., Waring BJ Simon JS (1993) Comparison of video thoracoscopic lung biopsy to open lung biopsy in the diagnosis of interstitial lung disease. Chest 103(3):765–770

20. Kreider ME, Hansen-Flaschen J, Ahmad NN et al (2007) Complications of video-assisted thoracoscopic lung biopsy in patients with interstitial lung disease. Ann Thorac Surg 83(3):1140–1144

21. Durheim MT, Kim S, Gulack BC et al (2017) Mortality and Respiratory Failure After Thoracoscopic Lung Biopsy for Interstitial Lung Disease. Ann Thorac Surg 104(2):465–470

22. Sheth JS, Belperio JA, Fishbein MC et al (2017) Utility of Transbronchial vs Surgical Lung Biopsy in the Diagnosis of Suspected Fibrotic Interstitial Lung Disease. Chest 151(2):389–399

23. Richardson CM, Pointon KS, Manhire AR Macfarlane JT (2002) Percutaneous lung biopsies: a survey of UK practice based on 5444 biopsies. Br J Radiol 75(897):731–735

24. Troy LK, Grainge C, Corte TJ et al (2020) Diagnostic accuracy of transbronchial lung cryobiopsy for interstitial lung disease diagnosis (COLDICE): a prospective, comparative study. The Lancet Respiratory Medicine 8(2):171–181

25. Herth FJ, Mayer M, Thiboutot J et al (2021) Safety and Performance of Transbronchial Cryobiopsy for Parenchymal Lung Lesions. Chest 160(4):1512–1519

26. Loor K, Culebras M, Sansano I et al (2023) Lung allograft transbronchial cryobiopsy for critical ventilated patients: a randomised trial. European Respiratory Journal 61(1):2102354

27. Ravaglia C, Wells AU, Tomassetti S et al (2019) Diagnostic yield and risk/benefit analysis of trans-bronchial lung cryobiopsy in diffuse parenchymal lung diseases: a large cohort of 699 patients. BMC Pulm Med 19(1):16

28. Zhao Y, Jiang F, Yu H et al (2021) Bronchus-blocked ultrasound-guided percutaneous transthoracic needle biopsy (BUS-PTNB) for intubated patients with severe lung diseases. Crit Care 25(1):359

29. Libby LJ, Gelbman BD, Altorki NK, Christos PJ Libby DM (2014) Surgical lung biopsy in adult respiratory distress syndrome: a meta-analysis. Ann Thorac Surg 98(4):1254–1260

30. Hashimoto H, Yamamoto S, Nakagawa H et al (2022) Clinical Utility of Surgical Lung Biopsy for Patients with Acute Respiratory Distress Syndrome: A Systematic Review and Meta-Analysis. Respiration 101(4):422–432

31. Dhooria S, Sehgal IS, Aggarwal AN, Behera D Agarwal R (2016) Diagnostic Yield and Safety of Cryoprobe Transbronchial Lung Biopsy in Diffuse Parenchymal Lung Diseases: Systematic Review and Meta-Analysis. Respir Care 61(5):700–712

32. Rodrigues I, Estêvão Gomes R, Coutinho LM et al (2022) Diagnostic yield and safety of transbronchial lung cryobiopsy and surgical lung biopsy in interstitial lung diseases: a systematic review and meta-analysis. Eur Respir Rev 31(166)

33. Wu CC, Maher MM Shepard JA (2011) Complications of CT-guided percutaneous needle biopsy of the chest: prevention and management. AJR Am J Roentgenol 196(6):W678–682

34. Gerard L, Bidoul T, Castanares-Zapatero D et al (2018) Open Lung Biopsy in Nonresolving Acute Respiratory Distress Syndrome Commonly Identifies Corticosteroid-Sensitive Pathologies, Associated With Better Outcome. Crit Care Med 46(6):907–914

35. Martin C, Papazian L, Payan MJ, Saux P Gouin F (1995) Pulmonary fibrosis correlates with outcome in adult respiratory distress syndrome. A study in mechanically ventilated patients. Chest 107(1):196–200

