## supplementary materials for "Safety and efficacy of bronchus-blocked ultrasound-guided percutaneous transthoracic needle biopsy (BUS-PTNB) for critically ill patients under invasive mechanical ventilation: a prospective, single-arm trial"

**CONTENTS** **Page**

Supplementary Figure 1 Workflow and role-based task assignments of BUS-PTNB……………..…..……2

Supplementary Figure 2 BUS-PTNB procedure images.…...………………....……….….………….……….……3

Supplementary Figure 3 CT and pathological images…………..….…………….....……….……………….……...4

**Supplementary Figure 1, Workflow and role-based task assignments of BUS-PTNB**

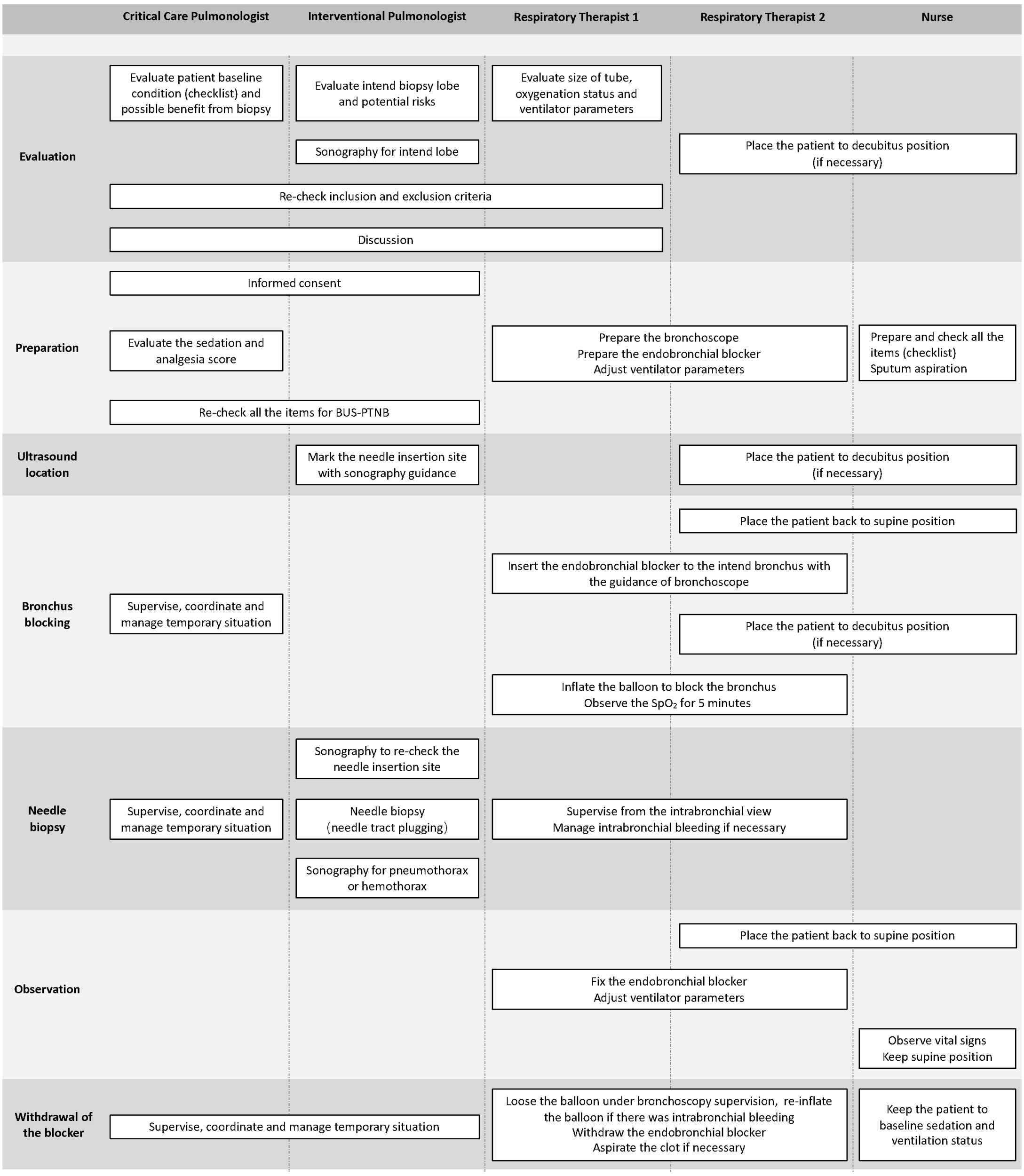


BUS-PTNB = bronchus-blocked ultrasound-guided percutaneous transthoracic needle biopsy, $SpO_2$ = oxygen saturation.

**Supplementary Figure 2, BUS-PTNB procedure images**

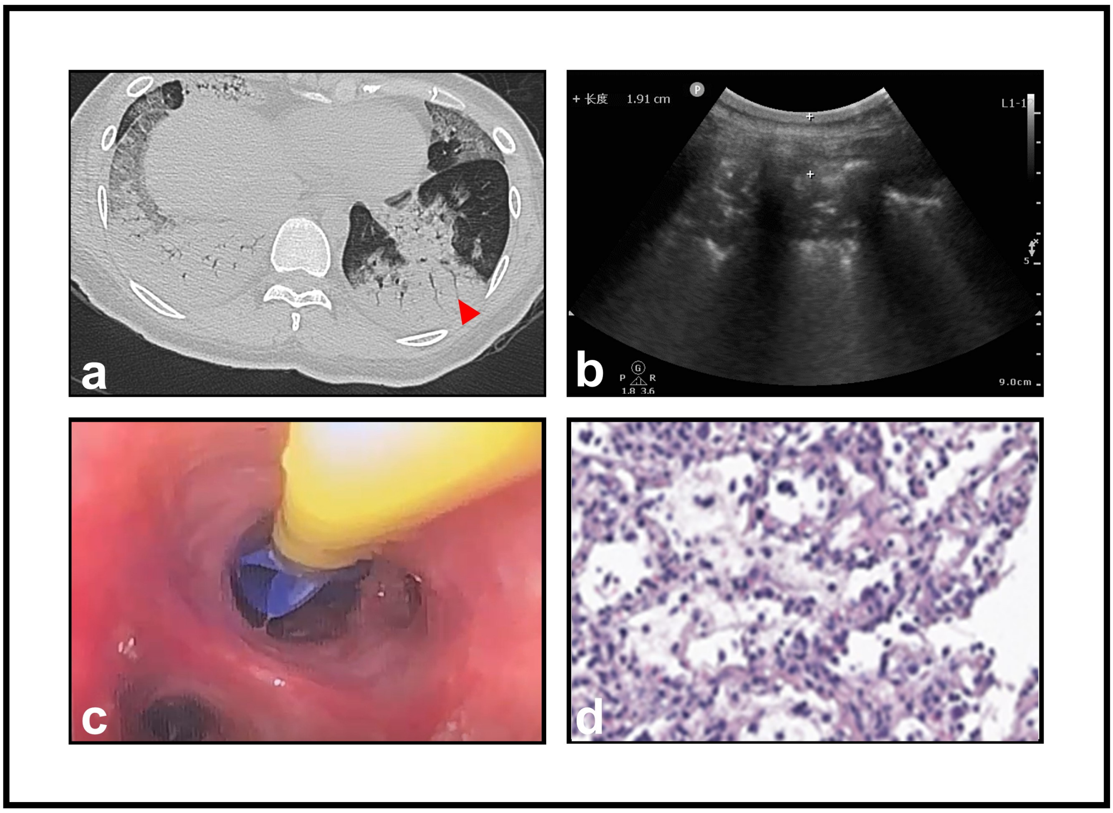


Participants were evaluated based on pre-procedural CT (a) and clinical conditions. Sonography was performed to confirm the exact puncture site at bedside (b). Then, the endobronchial blocker was placed into the intended lobar bronchus with a bronchoscope (c). Finally, tissue specimens were obtained using cutting needle and underwent pathological evaluation (d).

**Supplementary Figure 3, CT and pathological images**

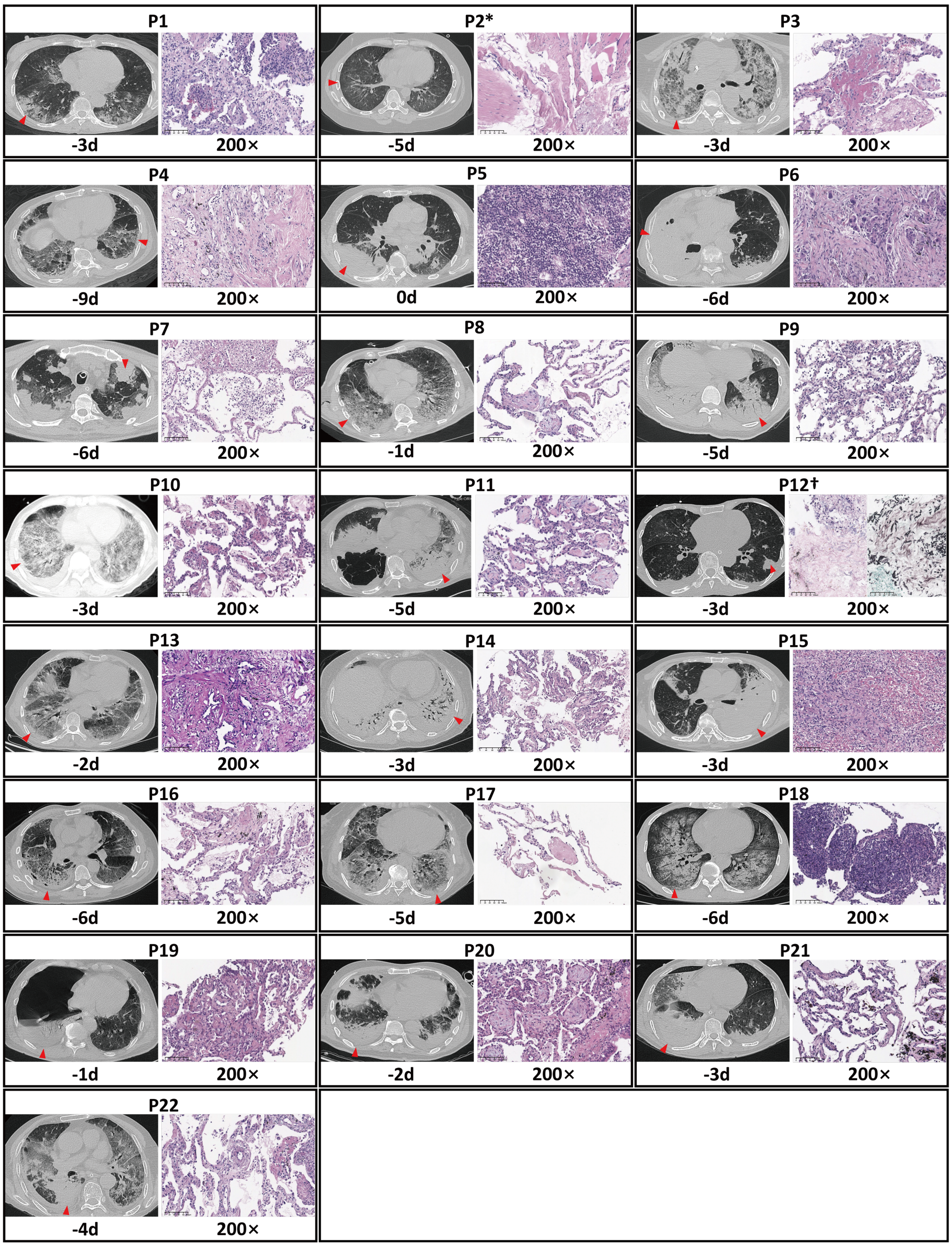


Biopsy sites and hematoxylin and eosin (H&E) staining of tissue specimens (N = 22). Red arrowheads indicate the sites of puncture. * No intended lesion. † Hyphae with parallel walls were observed in H&E and GMS staining, suggesting invasive aspergillosis.

CT = computerized tomography
